# Rare *ACTN2* Frameshift Variants Resulting in Protein Extension Cause Distal Myopathy and Hypertrophic Cardiomyopathy through Protein Aggregation

**DOI:** 10.1101/2024.01.17.23298671

**Authors:** Johanna Ranta-aho, Kevin J. Felice, Per Harald Jonson, Jaakko Sarparanta, Johanna Palmio, Giorgio Tasca, Mario Sabatelli, Cédric Yvorel, Ines Harzallah, Renaud Touraine, Lynn Pais, Christina A. Austin-Tse, Vijay Ganesh, Melanie C. O’Leary, Heidi L. Rehm, Michael K. Hehir, Sub Subramony, Qian Wu, Bjarne Udd, Marco Savarese

**Author notes:** <u>Corresponding author</u> Johanna Ranta-aho Folkhälsan Research Center Biomedicum 1, Haartmaninkatu 8, 00290, Helsinki, Finland. Marco Savarese and Bjarne Udd contributed equally to this study.

## Abstract

Distal myopathies are a group of rare, inherited muscular disorders characterized by progressive loss of muscle fibers that begins in the distal parts of arms and legs. Recently, variants in a new disease gene, *ACTN2*, have been shown to cause distal myopathy. *ACTN2*, a gene previously only associated with cardiomyopathies, encodes alpha-actinin-2, a protein expressed in both cardiac and skeletal sarcomeres. The primary function of alpha-actinin-2 is to link actin and titin to the sarcomere Z-disk. New *ACTN2* variants are continuously discovered, however, the clinical significance of many variants remains unknown. Thus, lack of clear genotype-phenotype correlations in *ACTN2*-related diseases, actininopathies, persists.

**Objective:** The objective of the study is to characterize the pathomechanisms underlying actininopathies.

**Methods:** Functional characterization in C2C12 cell models of several *ACTN2* variants is conducted, including frameshift and missense variants associated with dominant actininopathies. We assess the genotype-phenotype correlations of actininopathies using clinical data from several patients carrying these variants.

**Results:** The results show that the missense variants associated with a recessive form of actininopathy do not cause detectable alpha-actinin-2 aggregates in the cell model. Conversely, dominant frameshift variants causing a protein extension do produce alpha-actinin-2 aggregates.

**Interpretation:** The results suggest that alpha-actinin-2 aggregation is the disease mechanism underlying some dominant actininopathies, and thus we recommend that protein-extending frameshift variants in *ACTN2* should be classified as pathogenic. However, this mechanism is likely elicited by only a limited number of variants. Alternative functional characterization methods should be explored to further investigate other molecular mechanisms underlying actininopathies.

## 1 Introduction

*ACTN2* encodes alpha-actinin-2, a protein located in the sarcomeric Z-disk and expressed in both skeletal and cardiac muscles.^1^ The primary function of the protein is to bind actin and titin to the Z-disk, helping the sarcomere to maintain the structural integrity of its contractile apparatus. Additionally, it functions as a mechanosensor and a signaling hub for other Z-disk proteins.^2–5^

The alpha-actinin-2 protein is composed of an N-terminal actin-binding domain (ABD) containing two calponin homology domains (CH1 and CH2), a central rod with four spectrin-like repeats (SRs), and a calmodulin-like domain (CAMD) with two EF-hand motifs.^1, 3^ The ABD binds actin, while the CAMD binds to the N-terminal Z-repeats of titin.^6–8^ The interaction between titin and the CAMD is highly dynamic, and likely regulated by phospholipids and other intermolecular mechanisms.^3, 5, 9^

Variants in *ACTN2* are associated with a variety of muscle disease phenotypes, including both cardiac and skeletal muscle involvement, most notably cardiomyopathy and distal and congenital myopathy.^10–24^ Distal myopathies are a group of rare muscle disorders characterized by progressive loss of muscle tissue that primarily begins in the distal parts of arms and legs. In addition to *ACTN2*, more than 25 different genes have been associated with distal myopathy.^4^

Similarly, familial cardiomyopathies are a group of cardiac muscle disorders, defined by abnormal myocardial structure or function in the absence of other heart diseases or abnormal loading conditions. Cardiomyopathies can roughly be divided into five morphological subtypes: hypertrophic cardiomyopathy (HCM), arrythmogenic right ventricular cardiomyopathy (ARVC), dilated cardiomyopathy (DCM), restrictive cardiomyopathy (RCM), and left ventricular noncompaction (LVNC).^25, 26^ In relation to *ACTN2*, the clinical presentation is heterogenous, and indications of all types of cardiomyopathies have been documented in patients carrying *ACTN2* variants.^10–19^

While *ACTN2*-related diseases, actininopathies, are rare, *ACTN2* variants are identified in a growing number of patients. However, the interpretation of these variants is challenging, and genotype-phenotype correlations in actininopathies are poorly understood. The clinical presentation of distal myopathies in general is heterogenous, which is also true for actininopathies.^4, 27^ The severity, age of onset, and myopathological findings of the disease, as well as the distribution of affected muscle groups can vary significantly, which makes variant interpretation complex. Moreover, with a low number of patients, conclusive segregation analysis is difficult to achieve. Finally, only limited functional data to support variant pathogenicity is available.

It remains unclear why some variants result in an isolated cardiomyopathy, while others cause skeletal muscle disease or affect both muscle types. Functional studies characterizing the pathomechanisms underlying *ACTN2* variants have only been performed with a few variants. These studies suggest a few different disease mechanisms, mainly related to protein aggregation or altered interaction with other proteins.^12, 18, 22, 23^ However, the molecular mechanisms underlying most actininopathies remain poorly characterized.

Here, we investigate the pathomechanisms underlying actininopathies, and identify three novel frameshift variants. These variants cause adult-onset distal myopathy and/or cardiomyopathy, with or without facial weakness. We perform functional characterization *in vitro* to study the molecular consequences of *ACTN2* variants, providing insights to the molecular mechanisms underlying actininopathies.

## 2 Methods

### 2.1 Study Subjects and Clinical and Molecular Examinations

Three unrelated families were clinically and genetically investigated. Genomic DNA was isolated from blood cells using standard techniques. DNA of proband from family A was whole genome sequenced (WGS) at Genomics Platform at the Broad Institute of MIT and Harvard (Cambridge, MA, USA) using Illumina HiSeq X Ten v2 chemistry at a mean coverage of over 30x in the target region. WGS was performed on the DNA sample of proband from family B using 2X150bp read on Illumina next generation sequencing (NGS) system at a mean coverage of 40x in the target region (PerkinElmer Genomics, Pittsburgh, PA, USA).

DNA sample of proband from family C was sequenced using a targeted gene panel and the MiSeq Illumina system (San Diego, CA, USA).

Raw next generation sequencing (NGS) data were analyzed using a standard pipeline. *ACTN2* variants, described on the MANE transcript NM_001103.4, were confirmed by PCR and Sanger sequencing (primers available on request) and segregation analysis was performed on the available family members.

The recruited patients provided written informed consent to their referring clinician. The study was performed in accordance with the Declaration of Helsinki. Ethical approval was obtained through the institutional review board of Helsingin yliopistollinen sairaala – Hospital District of Helsinki and Uusimaa (HUS:195/13/03/00/11).

### 2.2 Muscle Histopathology

Right gastrocnemius muscle biopsy of a patient from Family B was processed and analyzed at the pathology lab at the University of Connecticut Health Center. Muscle tissue was obtained by open biopsy, frozen into the liquid phase of isopentane, previously cooled in liquid nitrogen. Transverse cryostat sections were cut at 10 μm thickness and stained by hematoxylin and eosin, modified Gomori trichrome, NADH-tetrazolium reductase (NADH-TR), succinate dehydrogenase, Congo red, periodic acid Schiff, cytochrome c oxidase according to standard protocols. Immunohistochemical analyses of muscle biopsy sections were performed with monoclonal primary antibodies against desmin, fast and slow myosin, dystrophin (rod, C-terminus and N-terminus domains), dysferlin, merosin, emerin, alpha- and gamma-sarcoglycan, and caveolin-3 using standard techniques. Samples for electron microscopy were fixed in 4% glutaraldehyde and processed according to standard procedures. Processed slides and electron microscopy photos were reviewed by an experienced neuromuscular medicine specialist (KJF) and neuropathologist (QW).

### 2.3 *ACTN2* Frameshift Variants

Several *ACTN2* frameshift variants have been previously observed in the last two exons of the gene (Table 1). In addition to the described patient variants, other variants were curated from literature and databases (LOVD, ClinVar; April 3^rd^, 2023). Both possible alternative reading frames are represented among the reported variants. The length of the altered protein sequence varies slightly between the variants. Some variants were observed in patients with skeletal (distal) myopathy, while others in patients with cardiomyopathy.

**Table 1.**
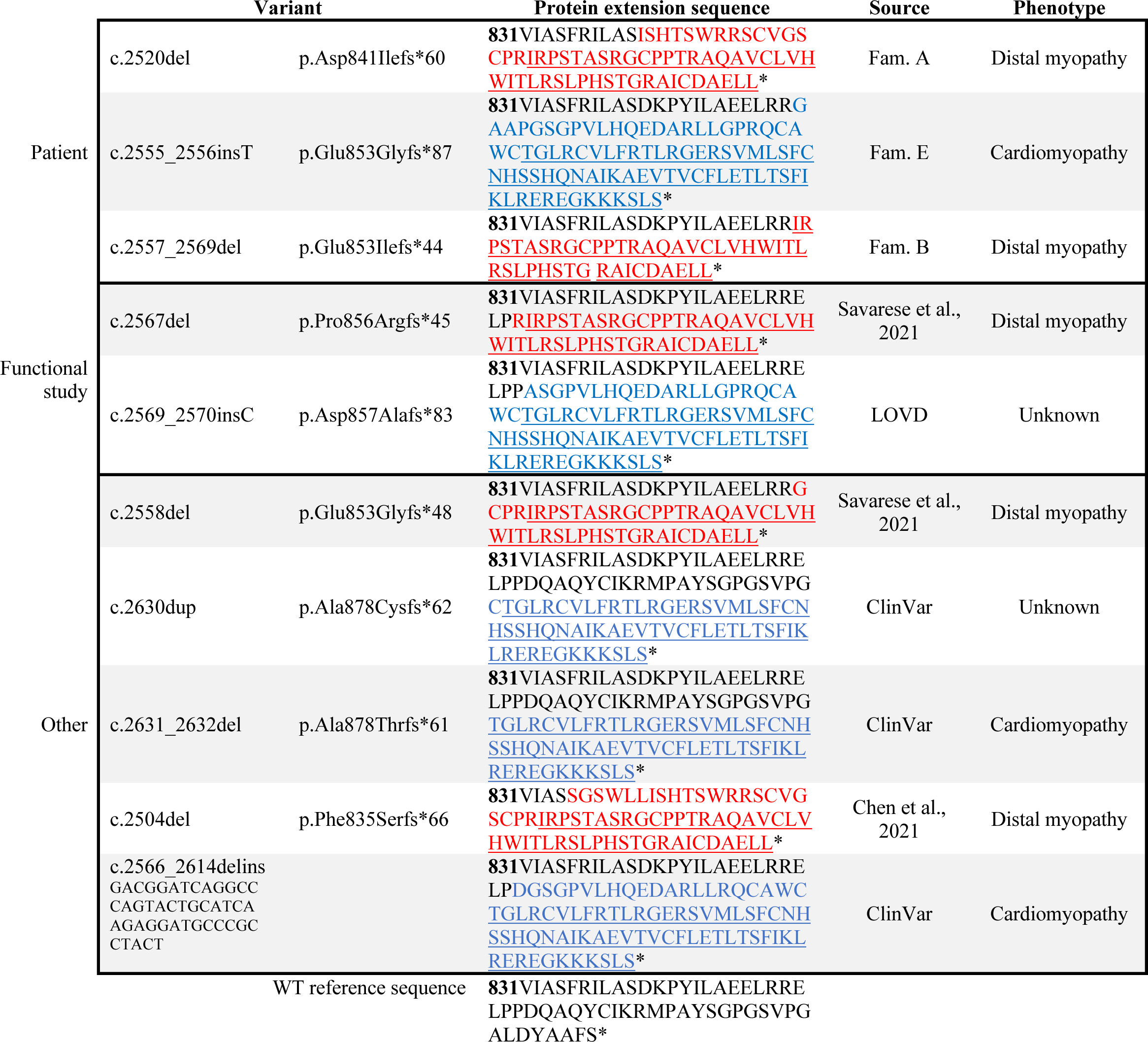
List of *ACTN2* frameshift variants identified in the described patients, and variants curated from literature and databases. The variants c.2567del and c.2569_2570insC were chosen for functional validation to represent the two alternative reading frames. All listed variants cause one of two protein extensions (distinguished by red and blue color) with slight variation in the length and starting point of the altered sequence. Mutant sequence shared between variants in the same reading frame is underlined.

### 2.4 Plasmid Constructs and Reagents

The wild-type construct was provided by Dr. Jocelyn Laporte. Recombinant eGFP-tagged mutated alpha-actinin plasmid constructs were generated by GenScript (New Jersey, USA). The mutant *ACTN2* constructs generated were c.2567del (p.Pro856Argfs*45), c.2569_2570insC (p.Asp857Alafs*83), and c.392T>C (p.Leu131Pro). The sequences of the plasmid constructs were verified by Sanger sequencing. Two mutant variants were chosen to represent each of the two possible alterative reading frames compared to the wild type. All known *ACTN2* frameshift variants are listed in Table 1, along with the predicted protein extension. The missense variant c.392T>C (p.Leu131Pro) was included as a positive control, as it has been previously shown to cause aggregation in a cell model.^9^

Additionally, several missense variant plasmid constructs were generated. These variants included c.1459T>C (p.Cys487Arg), c.2602G>A (p.Ala868Thr), c.893G>A (p.Arg298His), c.1070G>A (p.Arg357His), and c.2129A>G (p.Asn710Ser).

### 2.5 Cell Culture and Transfection

HeLa and C2C12 cells were maintained at 37°C, 5% CO_2_. HeLa cells were cultured in Minimum Essential Medium (MEM, Gibco, Massachusetts, USA) supplemented with 10% fetal bovine serum (FBS) (Thermo Fisher Scientific, Massachusetts, USA), 1% GlutaMax (Gibco), and 1% penicillin/streptomycin (PS) (Thermo Fisher Scientific). C2C12 cells were maintained in Dulbecco’s modified Eagle Medium (DMEM, Gibco) supplemented with 20% FBS, 1% GlutaMax, and 1% PS.

C2C12 cells were cultured on either collagen-coated plasticware or on ultra-compliant gelatin hydrogel. Collagen coating was performed in collagen concentration of 0.05 mg/ml (Collagen type 1, from bovine, Thermo Fisher Scientific) in 20 mM acetic acid for 1 hour. Gelatin hydrogel coating was prepared with 2.5% porcine skin gelatin (G2625, Sigma-Aldrich, Missouri, USA) and ∼10 U/ml microbial transglutaminase (Probind TX, BDF Natural Ingredients S.L., Spain) in PBS.^28^

For myotube differentiation, myoblasts were grown to confluency on hydrogel-coated plasticware and cultured in differentiation medium (DMEM with 2% heat-inactivated horse serum (Gibco), L-glutamine (Gibco), PS, and 10% OPTI-MEM (Thermo Fisher Scientific)). The cells were differentiated for up to 21 days.

HeLa cells were transfected with FuGENE 6 (Promega, Wisconsin, USA) and C2C12 cells with Lipofectamine 3000 (Invitrogen, Massachusetts, USA) according to manufacturer’s instructions.

### 2.6 Immunofluorescence Analysis

C2C12 myoblasts on collagen-coated plates were fixed 24 hours after transfection, while myotubes cultured on hydrogel-coated plates were fixed after the 21-day differentiation. All cells were fixed in 4% paraformaldehyde for 15 minutes. Fluorescence images were captured by Zeiss AxioCam 503 system (software Zeiss Zen 2.3, Carl Zeiss Microscopy GmbH, Germany) or EVOS FL system (Thermo Fisher Scientific).

### 2.7 Protein Solubility Assay

Cells were pelleted and lysed with lysis buffer composed of 150 mM NaCl, 10 mM tris-HCl (pH 7.5), 1% Halt Protease Inhibitor Cocktail (Thermo Scientific), 1% Triton X-100, and ∼ 25 U/ml Pierce Universal Nuclease for Cell Lysis (Thermo Scientific) for 1 hour at 4 °C. Trituration was performed through a 27-gauge needle during the lysis period. Subsequent centrifugation at 16000 g (Biofuge Fresco, Heraeus Instruments, Germany) resulted in a soluble (supernatant) and insoluble (pellet) fraction. The pellet was resuspended in a wash buffer containing 150 mM NaCl, 10 mM tris-HCl (pH 7.5), 1% Triton X-100, and centrifuged as above. Both soluble and insoluble fractions were Western blotted, and proteins were detected by monoclonal mouse anti-GFP antibody (6602-1-Ig, Proteintech, Illinois, USA) and donkey anti-mouse IgG (Invitrogen Alexa Fluor Plus 800) on the Odyssey imager (Li-Cor, Lincoln, NE, USA).

### 2.8 Statistical Analysis

Fiji software version 2.3.0/1.53q^29^ was used for data analysis and band quantification, statistical analysis was performed with IBM SPSS Statistics software (version 28.0.0.0, IBM corp., 2021). All experiments were repeated at least three times independently. Kruskal-Wallis *H* test was performed, followed by Dunn’s test for pairwise comparisons.

## 3 Results

### 3.1 Clinical Features and Genetic Findings

#### Family A

The proband was found to carry two *ACTN2* variants in cis: c.2194G>A (p.Ala732Thr) and c.2520del (p.Asp841Ilefs*60). Both variants were also identified in three symptomatic family members and were absent from the proband’s asymptomatic parent. The proband and the symptomatic parents presented with distal and proximal muscle weakness, with the parent also presenting with cardiomyopathy. Two other relatives also presented with left ventricular non-compaction cardiomyopathy.

#### Family B

WGS identified a heterozygous *ACTN2* c.2557_2569del (p.Glu853Ilefs*44) variant in the proband, and targeted DNA testing identified the same variant in five additional affected family members. The proband presented with distal and mild proximal weakness, as wells as with mild lower facial weakness. The proband’s two children presented with distal and lower facial weakness. One of the children also showed proximal weakness, while the other presented with atrial fibrillation and septal defect and had an enlarged left atrium. The proband’s sibling and two cousins all presented with distal and proximal weakness. The sibling also presented with atrial fibrillation and septal defect, while one of the cousins showed atrio-ventricular and right bundle branch block with mild diastolic dysfunction. One of the probands parents, grandparents, and great-grandparents all had leg weakness and gait difficulties, but they were all deceased and not available for further studies.

#### Family C

*ACTN2* c.2555_2556insT (p.Glu853Glyfs*87) variant was identified in the proband. The patient presented with atrial fibrillation and hypertrophic cardiomyopathy. There was no family history of cardiac disease.

MRI and histopathological findings from Family B are shown in Figure 1. Full clinical findings and pedigrees are available upon request.

**Figure 1.**
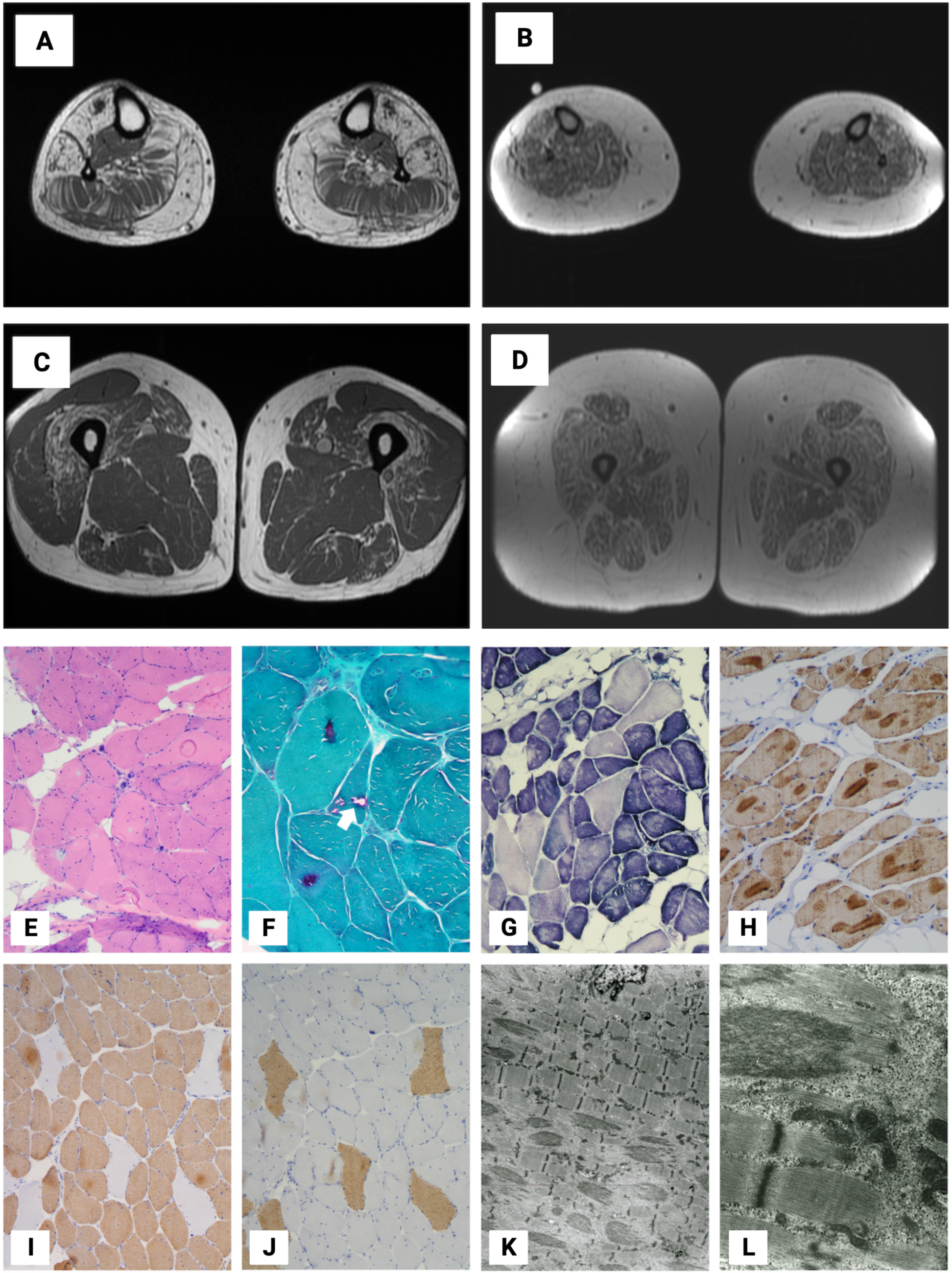
**A-D** Muscle MRI studies of family B. **A** Foreleg of proband’s sibling shows severe fatty atrophy of the anterior tibialis, peroneus and medial head of the gastrocnemius; moderate fatty atrophy of the soleus; and mild fatty atrophy of the distal portion of the posterior tibialis. **B** Foreleg of proband shows diffuse bilateral muscle atrophy, mild fatty atrophy of the vastus medialis, and mild subcutaneous edema along the anterolateral aspect of the lower legs without organized fluid collection or mass. **C** Thigh of proband’s sibling shows moderate-to-severe fatty atrophy of the vastus intermedius muscle; and mild fatty atrophy of the proximal vastus lateralis, tensor fascia lata, sartorius, biceps femoris, semimembranosus, and gracilis. **D** Proband’s thigh shows diffuse bilateral muscle atrophy. **E-L** Muscle histopathology of the proband’s sibling of family B. **E** Increased endomysial fatty infiltration, variation in myofiber size and internalized nuclei (H&E). **F** Rimmed vacuoles (arrow) (Gomori trichrome. **G** Cores (NADH-TR). **H** Increased desmin-reactivity. **I-J** Predominance and mild atrophy of type 1 myofibers (myosin slow and fast). **K-L** Focal filamentous disruption of sarcomeres (electron microscopy).

### 3.4 Alpha-actinin-2 Subcellular Localization and Solubility

To investigate the biological effects of the novel *ACTN2* frameshift variants, eGFP-tagged plasmid constructs c.2567del (p.Pro856Argfs*45), c.2569_2570insC (p.Asp857Alafs*83), c.392T>C (p.Leu131Pro), along with wild-type *ACTN2*, were generated. Each of these mutant frameshift constructs represents one of the two possible alternative reading frames. The missense variant c.392T>C (p.Leu131Pro), previously shown to produce protein aggregates,^23^ acted as the positive control. At both stages of differentiation (myoblast and myotube) transfected C2C12 cells expressing wild-type eGFP-ACTN2 showed a uniformly dispersed localization of the protein, whereas cells expressing one of the mutant constructs were characterized by protein aggregates in the cytosol (Figure 2). The two frameshift constructs resulted in protein aggregation indistinguishable from each other.

**Figure 2.**
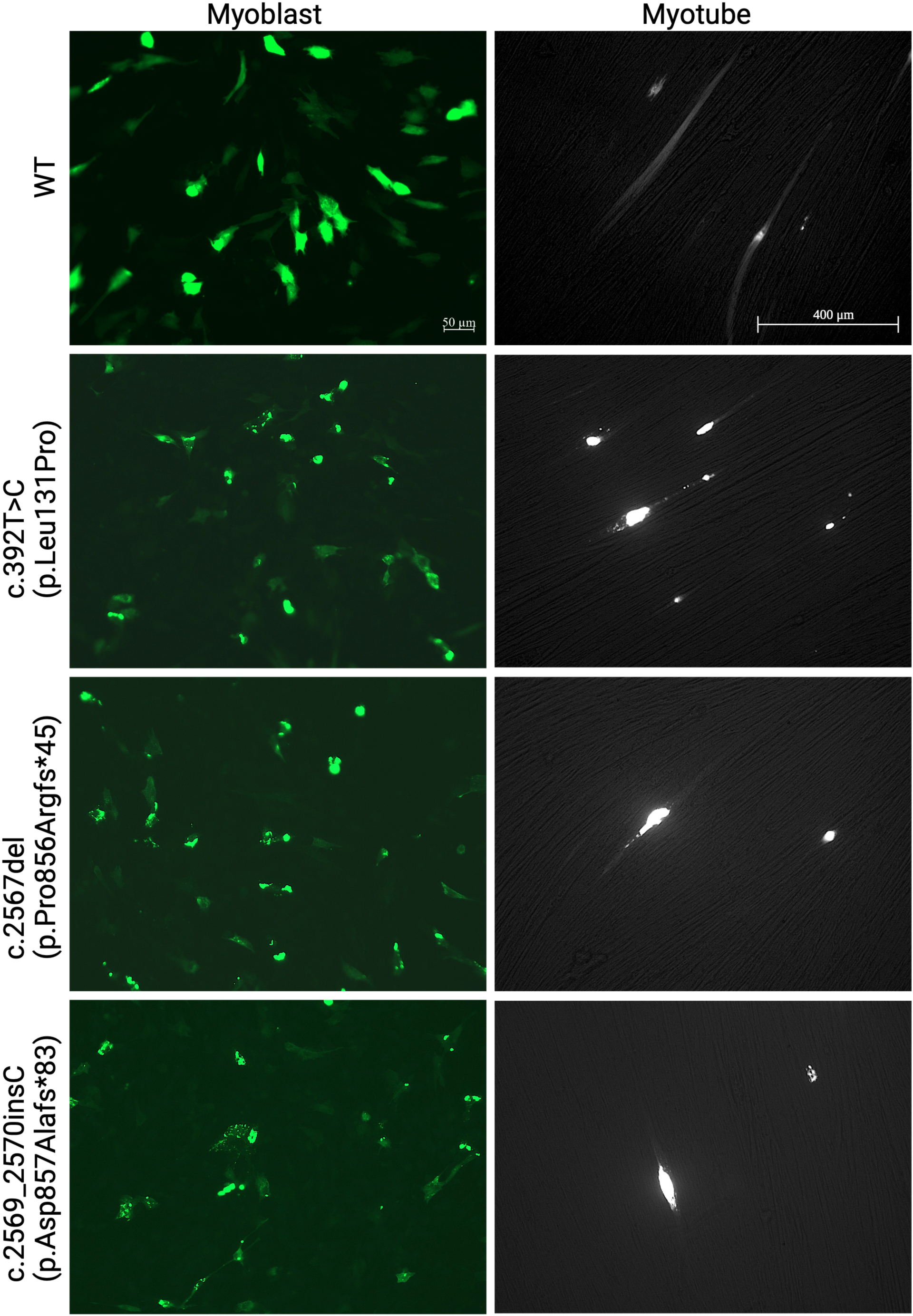
Alpha-actinin-2 localization in C2C12 myoblasts and myotubes expressing WT or mutant eGFP-ACTN2.

The presence of alpha-actinin-2 aggregation was further investigated through the solubility assay. In transfected HeLa cells, the wild-type eGFP-ACTN2 construct was detected in both insoluble and soluble fractions, whereas the frameshift mutant construct and c.392T>C (p.Leu131Pro) were almost exclusively present in the insoluble fraction (Figure 3). This result was consistent with the observed aggregates in C2C12 cells transfected with the mutant constructs and confirmed that the frameshift variants c.2567del (p.Pro856Argfs*45) and c.2569_2570insC (p.Asp857Alafs*83) cause alpha-actinin-2 aggregates in the cell.

**Figure 3.**
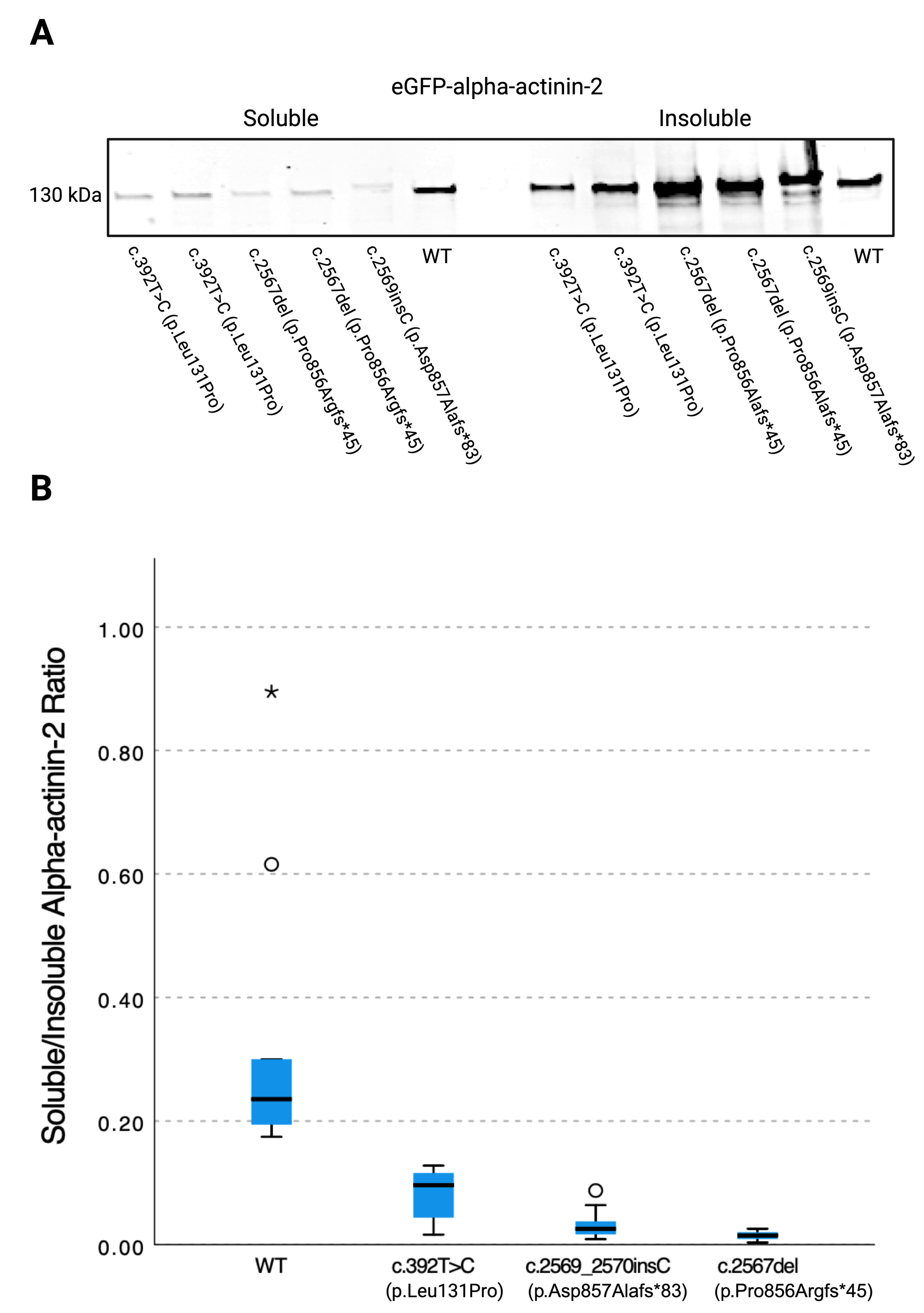
**A** Soluble and insoluble protein fraction per variant with western blot probing for eGFP-alpha-actinin-2. **B** eGFP-alpha-actinin-2 in the soluble versus insoluble protein fraction, shown as soluble over insoluble ratio. °mild outlier, *extreme outlier, bars denote minimum and maximum non-outlier values. n = 3 biological replicates.

The same experiment, including both the immunofluorescence microscopy and solubility assay, was also performed with various *ACTN2* missense variants. Variants included in the experiment were c.1459T>C (p.Cys478Arg)^24^, 2602G>A (p.Ala868Thr), c.893G>A (p.Arg298His), c.1070G>A (p.Arg357His), and c.2129A>G (p.Ans710Ser). However, none of these variants resulted in protein aggregation (not shown). Thus, c.392T>C (p.Leu131Pro)^24^, characterized by Chen and colleagues,^23^ still remains the only known protein aggregation-causing missense variant in *ACTN2*.

## 4 Discussion

*ACTN2* variants are known to cause a wide variety of skeletal muscle and cardiac phenotypes, making their clinical interpretation challenging.^10–24, 30^ Further, limited functional data is available for most *ACTN2* variants, and thus, clinical significance of many variants remains uncertain.^27^ Consequently, many variants are still classified as variants of uncertain significance (VUS), according to the American College of Medical Genetics / Association for Molecular Pathology (ACMG/AMP) guidelines.^31^ Almost all *ACTN2* frameshift variants included in this study were classified as VUS according to the guidelines, including our patient variants. Thus, the patients do not have a formal diagnosis of actininopathy.

Our study showed that protein-extending *ACTN2* frameshift variants c.2567del (p.Pro856Argfs*45) and c.2569_2570insC (p.Asp857Alafs*83) cause protein aggregation, suggesting that the protein misfolds and causes a dominant toxic effect on the cellular level. These variants are expected to escape nonsense-mediated decay (NMD)^32^ and produce an extended protein, as the variants are located in the last two exons and the new stop codon is predicted to fall in the 3’ untranslated region (3’-UTR). Thus, the result is a protein extension. Variants producing extended proteins in both possible alternative reading frames were identified. This study showed that variants representing both reading frames produce aggregation in the cell model, and a third variant (also listed in Table 1) has previously been found to do the same.^23^ As the deleted amino acid stretch is of similar length and the altered protein sequences are very similar to each other within the same reading frame, it is reasonable to assume that all currently known protein-extending variants downstream of the NMD site in *ACTN2* lead to protein aggregation. Thus, this study identified a class of *ACTN2* variants with a clear toxic gain-of-function pathomechanism. Additionally, variants belonging to this category were found to segregate with the diseases in families A and B. Considering the functional and segregation evidence outlined in this study, we suggest that the frameshift variants identified in families A, B, and C are disease-causing.

Currently, according to the ACMG/AMP guidelines,^31^ the variants identified in families A, B, and C (c.2520del, c.2557_2569del, and c.2555_2556insT, respectively) are interpreted as VUS, as PM2_Supporting and PVS1_Moderate criteria are be applied. Considering the functional and segregation evidence outlined in this study, we suggest that additional criteria should be applied. PS3_Moderate, and PP1_Supporting should be applied to variants identified in families A and B, changing the classification to likely pathogenic. Similarly, PS3_Moderate, PVS1_Moderate, PM2_Supporting should be applied to the variant identified in family C, raising the classification to likely pathogenic.

In family A, the protein-extending frameshift variant was identified in *cis* with another *ACTN2* missense variant c.2194G>A (p.Ala732Thr). While we cannot exclude a synergic effect of these two variants in family A, we expect, based on our functional data, that the frameshift variant would still be disease-causing by itself.

This study demonstrated that protein-extending frameshift variants in *ACTN2*, along with the previously characterized missense variant c.392T>C (p.Leu131Pro),^23^ cause protein aggregation *in vitro*. However, this is not true for all reported missense variants in *ACTN2*. No such effect was found in the variant c.2180T>G (p.Leu727Arg).^23^ In this study, we found that also c.1459T>C (p.Cys487Arg) (not shown) associated with a dominant actininopathy, does not cause aggregation in the cell models. Thus, we suggest that multiple disease mechanisms are involved in dominant actininopathies. However, these alternative mechanisms still remain poorly characterized. As c.2180T>G (p.Leu727Arg) is located in close proximity to the CAMD, which binds to the N-terminal Z-repeats of titin, we hypothesize that the pathomechanism may be related to alterations in this interaction. Similarly, variants located in the ABD may cause the disease through alterations in the binding of alpha-actinin-2 with actin.^12, 16, 33, 34^ However, more research is required to uncover these alternative disease mechanisms.

Clinically, family A includes individuals with a strict skeletal muscle disease, individuals with a strict cardiac phenotype, and an individual with both. In family B, all affected individuals present with skeletal muscle weakness, some of them showing cardiac involvement as well. On the other hand, the proband of family C presents with a strict cardiac phenotype, without any family history of muscle or cardiac disease. It is not clearly understood why such varying phenotypes are observed within a single family, or more generally, why some *ACTN2* variants result in an isolated cardiomyopathy, while others cause a skeletal muscle disease.^27^ Prior to this study, only a few patients with skeletal muscle actininopathy have been observed with an overt cardiac involvement.^20, 23^ It is possible that in patients with a cardiac actininopathy, mild skeletal muscle weakness maybe overlooked. It may also be possible that the two tissue types become affected at different stages of the disease. In other words, younger patients with cardiomyopathy symptoms may develop a skeletal muscle phenotype at a later stage of the disease, or vice versa. Biologically, a modifying gene may be involved in the determination of the affected tissue type.

## 5 Conclusion

The identification of protein-extending frameshift variants in three independent families and the presented functional data strongly supporting their pathogenicity outlines a new category of pathogenic variants in *ACTN2*. This finding will aid variant interpretation in the future, and it is a step forward in the characterization of the pathomechanisms underlying actininopathies. Clinically, the finding will have an immediate impact, as it will allow patients carrying these variants to receive a final diagnosis and accurate genetic counseling.

## 5 Acknowledgements

We would like to thank Merja Soininen and Helena Luque for technical assistance. The work was supported by the Folkhälsan Research Foundation (to MS and BU), the Jane and Aatos Erkko Foundation (BU), the Sigrid Jusélius Foundation (MS and BU), Finska Läkaresällskapet (BU), Association Française contre les Myopathies (grant #23281 to MS), Academy of Finland (grant #339437 to MS), European Joint Programme for Rare Disease (BU), and Sydäntutkimussäätiö (MS). Sequencing and analysis of Family A were provided by the Broad Institute of MIT and Harvard Center for Mendelian Genomics (Broad CMG) and were funded by the National Human Genome Research Institute grants UM1 HG00890 (with additional support from the National Eye Institute, and the National Heart, Lung and Blood Institute), U01 HG0011755, and R01 HG00914 and in part by CZI grant DAF2019-199278 and grant DOI https://doi.org/10.37921/236582yuakxy from the Chan Zuckerberg Initiative DAF, an advised fund of Silicon Valley Community Foundation (funder DOI 10.13039/100014989).

## 6 Author Contributions

Conception and design of the study: JR, M. Savarese, PHJ, JS, BU; Acquisition and analysis of data: JR, PHJ, JS, JP, GT, M. Savarese, CY, IH, RT, LP, CAT, VG, MO, HR, MKH, SS, QW, KJF, M. Sabatelli; Drafting of the manuscript: JR, M. Savarese, BU, MKH, SS, QW, KJF, LP, MO, RT

## 8 Data Availability

The data that supports the findings of this study are being shared or are available in Leiden Open Variation Database at https://databases.lovd.nl/shared/genes/ACTN2 and in ClinVar (National Library of Medicine, National Institute of Health) https://www.ncbi.nlm.nih.gov/clinvar/?term=ACTN2[gene].

